# Impact of natural disasters on HIV risk behaviors, seroprevalence, and virological supression in a hyperendemic fishing village in Uganda

**DOI:** 10.1101/2023.10.19.23297262

**Authors:** Hadijja Nakawooya, Victor Ssempijja, Anthony Ndyanabo, Ping Teresa Yeh, Larry W. Chang, Maria J. Wawer, Fred Nalugoda, David Serwadda, Ronald H. Gray, Joseph Kagaayi, Steven J Reynolds, Tom Lutalo, Godfrey Kigozi, M. Kate Grabowski, Robert Ssekubugu

## Abstract

**Background:** Understanding the impact of natural disasters on the HIV epidemic in populations with high HIV burden is critical for the effective delivery of HIV control efforts. We assessed HIV risk behaviors, seroprevalence, and viral suppression in a high-HIV prevalence Lake Victoria fishing community before and after COVID-19 emergence/lockdown and a severe lake flooding event, both of which occurred in 2020.

**Methods:** We used data from the largest Lake Victoria fishing community in the Rakai Community Cohort Study, an open population-based HIV surveillance cohort in south-central Uganda, collected prior to (September-December 2018) and after (October-December 2021) COVID-19 emergence/lockdown and a severe flooding event, to evaluate the impact of natural disasters on the key population-level HIV outcomes listed above. Households impacted by flooding were identified using drone data and through consultation with village community health workers. The entire study population was subject to extensive COVID-19-related lockdowns in the first half of 2020. Differences in HIV-related outcomes before and after COVID, and between residents of flooded and non-flooded households, were assessed using a difference-in-difference statistical modeling approach.

**Findings:** 1,226 people participated in the pre- and post-COVID surveys, of whom 506 (41%) were affected by flooding and 513 (41%) were female. HIV seroprevalence in the initial period was 37% in flooded and 36.8% in non-flooded households. Following the COVID-19 pandemic/lockdown, we observed a decline in HIV-associated risk behaviors. Transactional sex declined from 29.4% to 24.8% (p=0.011), and inconsistent condom use with non-marital partners declined from 41.6% to 37% (p=0.021) in the pre- and post-COVID periods. ART coverage increased from 91.6% to 97.2% (p<0.001). There was 17% decline in transactional sex (aPR=0.83, 95% CI: 0.75-0.92) and 28% decline in the overall HIV risk score (aPR=0.83, 95% CI: 0.75-0.92) among HIV-seronegatives and 5% increase in ART coverage between the pre- and post-COVID periods. We observed no statistically significant differences in changes of HIV risk behavior, seroprevalence, or viral suppression outcomes comparing those affected by floods to those not affected by floods in the periods before and after COVID in difference-in-difference analyses.

**Interpretation:** Despite a high background burden of HIV, the COVID-19 pandemic, and severe flooding, we observed no adverse impact on HIV risk behaviors, seroprevalence, or virologic outcomes. This may be attributed to innovative HIV programming during the period and or population resilience. Understanding exactly what HIV programs and personal/community-level strategies worked to maintain good public health outcomes despite extreme environmental and pandemic conditions may help improve HIV epidemic control during future natural disaster events.

## INTRODUCTION

Globally, natural disasters have grown in frequency and severity, with flooding being among the most common and devastating disaster events[1, 2]. Over the last decade, African countries have been especially hard hit by flooding, with climate change driving recent, large increases in flooding events[3-5]. The African continent is also home to the largest number of HIV cases globally [6]. Prior research suggests that floods can negatively impact HIV epidemic control due to interruptions in HIV service provision and changes in human behaviors [7-9].

Lake Victoria fishing communities in eastern Africa have one of the highest HIV burdens globally, with adult HIV seroprevalence estimates ranging from 20 to 40% [10, 11]. Since early 2020, residents along the shores of Lake Victoria in East Africa have been experiencing the health, social, and economic impacts of severe flooding events as well as the COVID-19 pandemic in the early 2020s [12-15], raising concern that hard-earned gains in HIV epidemic control made over the prior decade in these high-burden settings might be lost [16-18].

In Uganda, people living with HIV have experienced extreme stress due to the COVID-19 pandemic, including worries about access to antiretroviral therapy (ART), concern over inadvertent disclosure of HIV status, fear that coronavirus infection would have more severe outcomes for people living with HIV (PLHIV), and distress due to pandemic-induced poverty and food insecurity [16]. Persons living along the Lake Victoria shoreline experienced added stresses due to lake flooding events following heavy continuous rains [19]. As a result of flooding, communities along the shorelines were grossly affected in terms of their livelihood including displacement of families and households [20]. Despite extremely high HIV burden in Lake Victoria fishing communities, it is largely unknown how these dual natural disasters have impacted the state of the HIV epidemic, including possible effects on population HIV-associated risk behaviors and treatment outcomes among PLHIV.

Using data from the Rakai Community Cohort Study (RCCS), an open longitudinal population-based HIV surveillance cohort, we conducted and pre and post study to evaluate the impact of COVID-19 and flooding on the prevalence of HIV-associated risk behaviors and HIV virologic suppression among PLHIV. Our analysis included data from individuals residing in a large Lake Victoria fishing community that experienced severe flooding in 2020, during the earlier times of COVID-19 pandemic national lockdowns. HIV burden within this community ranks among the highest in the East African region, with an estimated incidence rate of ∼1.6/100 per person years and an HIV seroprevalence of 37% [21]. Specifically, we examined changes in HIV-associated behaviors and treatment outcomes before and after COVID-19 among individuals who resided in the community affected by severe flooding and COVID-19 emergence. Research on health outcomes following natural disasters may help inform public health policies to prevent and reduce negative health outcomes during times of emergency.

## METHODS

### Study design and participants

The RCCS is an open population-based longitudinal HIV surveillance cohort in south central Uganda, that is implemented in agrarian, semi-urban and four Lake Victoria fishing communities. The RCCS has been ongoing in agrarian and semi-urban trading communities since 1994 and Lake Victoria fishing communities since 2011, and has been described in detail elsewhere [11, 22]. Briefly, eligible participants are identified through a household census enumeration of all person’s information on household members’ sex, age, and duration of residence obtained regardless of whether they are present in the home at time of census. During census, household geographic coordinates (latitude and longitude) are collected using global positioning system (GPS) technologies, using the Garmin eTrex 10 GPS. After the census, the RCCS surveys all residents within the eligible age range (15-49 years) who can provide written informed consent. Participants are then interviewed regarding demographic characteristics, sexual behaviors, and ART use. Venous blood is obtained for HIV testing, and viral assessment was conducted among HIV-positive persons.

This study included the RCCS participants residing in the four RCCS fishing communities, and the largest Lake Victoria fishing community in south central Uganda [21]. Analyses were restricted to two RCCS survey rounds (Figure 1). The pre-study period included the last RCCS survey done prior to the onset of the COVID-19 pandemic in March 2020 and Lake Victoria flooding in February 2020 (conducted between September 3rd, 2018, and December 19^th^, 2018). The post-study period included the first RCCS survey conducted after the onset of the pandemic and flooding (conducted between October 11^th^, 2021, and December 16^th^, 2021). Because of widespread migration during the period of analysis, we used longitudinal analysis to limit the analysis to only individuals who participated in both RCCS surveys and thus were likely to have been exposed to the two emergencies.

**Figure 1:**
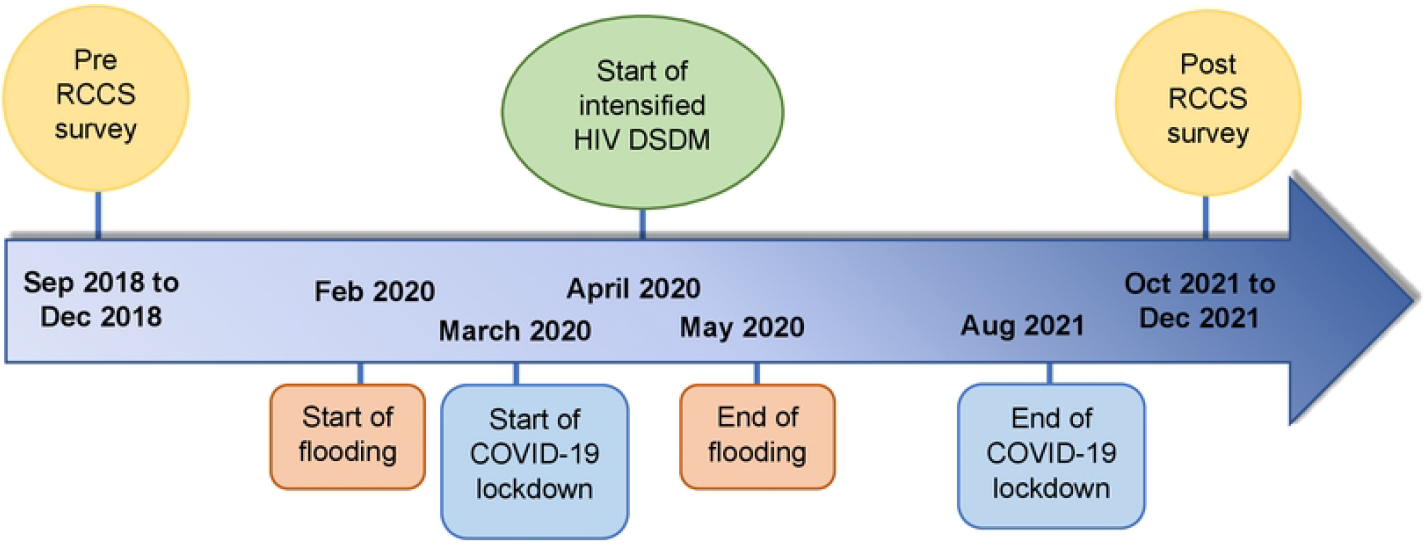
Timeline of key events, including timing of RCCS surveys, Lake Victoria flooding, COVID-19 emergence, intensified implementation of differentiated HIV service delivery models (DSDM).

### Provision of HIV services in fishing communities during the COVID-19 pandemic and Lake Victoria flooding

The adaptation of HIV services in fishing communities during COVID-19 in Uganda were based on guidelines provided by the Uganda Ministry of Health to all HIV service implementing partners throughout the country. The different models that were used to accommodate COVID-19 restrictions have been described elsewhere [23]. In general, multiple community-based strategies were used to ensure continued access to essential HIV services during lockdowns and travel restrictions. For example, community drug dispensing points (CDDP) were set up: health workers moved to communities to meet with clients and deliver their drugs. Four CDDP sites were operated in this fishing village (including one on a nearby island), and each of these sites was visited at least once every month. In addition, under the community client-led ART delivery model, clients within the same geographical location formed homogenous groups of not more than ten people, in which the group leader (or designated member) would travel to the CDDP or health facility to collect drugs for the rest of the members, to decrease congestion during drug pickup. Within all the models of differentiated services, multi-month dispensing of drugs was encouraged; stable clients would be given drug of up to six monthly drug refills and all this was supplemented by mobile phone counseling and check-in calls.

### Identification of flooded households

Flooding happened continuously for six months following heavy rains that started in February 2020 [24]. The floods submerged extended distances of the shores of Lake Victoria as a result some households were submerged in the water, got washed away, or became grossly damaged and/or uninhabitable. To identify affected households, a drone-supported GPS was launched to identify the new shoreline of Lake Victoria following the flooding to determine which households were affected by the floods. By capturing coordinates of the shoreline, it was possible to compare these with the previous shoreline to see which households were located within the flooded area along the lake. This exercise was conducted before the shoreline receded to its original position. We also identified other offshore patches that were affected by the floods through consultation with village community health workers and local leaders who were present at the time of floodings. After identification of the offshore flooded areas, the drone was used to map and capture coordinates for the off-shoreline patchy areas as well. We plotted all active clusters of households that existed before flooding using ARCGIS-PRO and fitted the current shoreline and the offshore map of the flooded area. All households below the shoreline or in the flooded map were identified as flooded and the rest were identified as non-flooded (Figure 2). We also assessed changes in household location after flooding to identify new areas of settlement.

**Figure 2:**
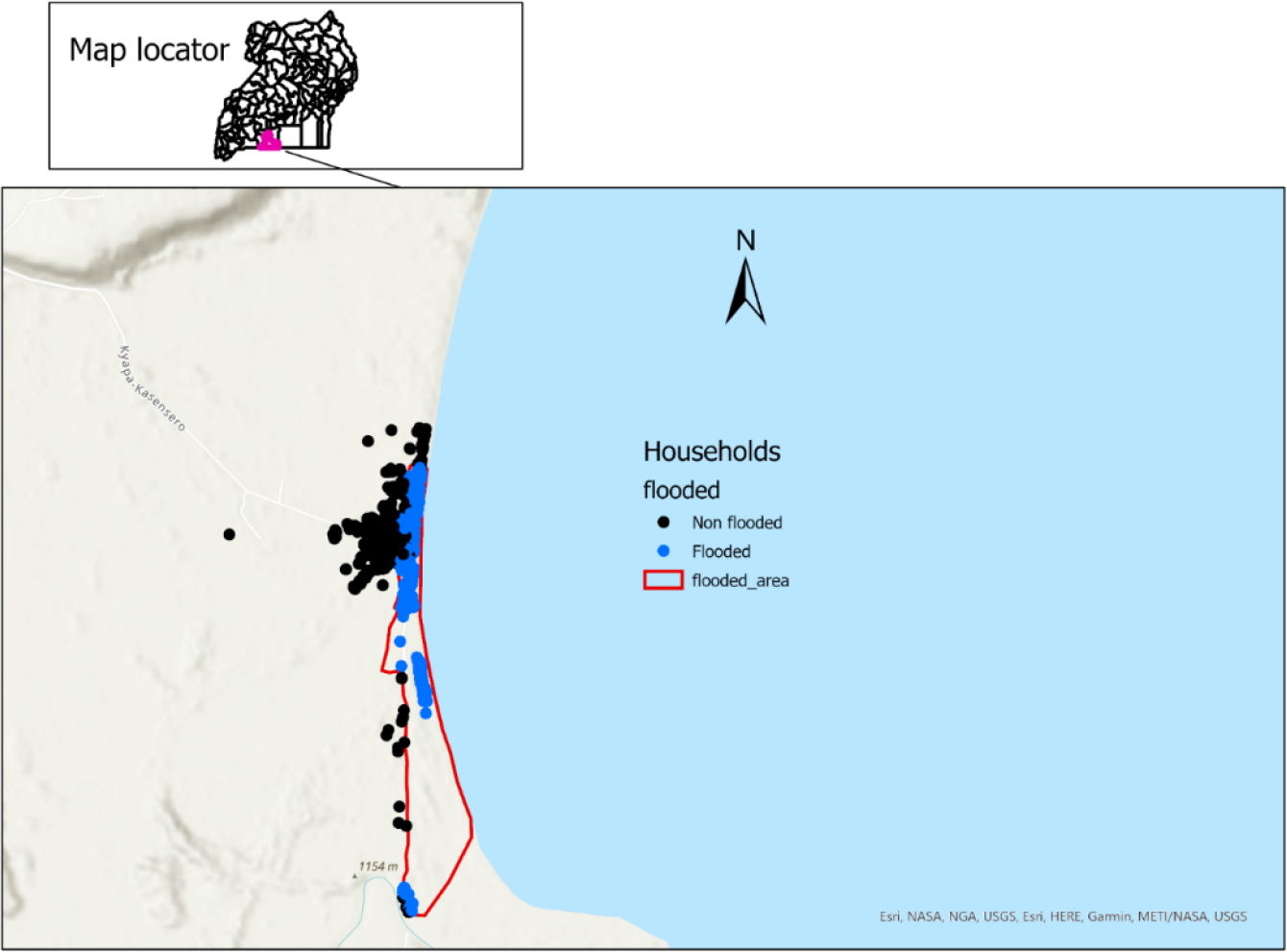
Map of flooded households following a mass flooding event in spring of 2020 in Kasensero, Uganda

### Primary outcomes

Primary HIV-associated behaviors and treatment outcomes included number of sexual partners in the past 12 months, inconsistent condom use with non-marital partners, transactional sex, alcohol use in the past 12 months, HIV prevalence, ART use, and viral suppression. These were assessed before and after flooding in pre-COVID-19 and post-COVID-19 RCCS surveys. HIV risk score among HIV-seronegative participants was measured using a previously validated gender-specific risk index to predict risk of HIV infection [25]. Individuals were assigned a risk score using the following variables: age, marital status, education, number of sexual partners, frequency of condom use, use of alcohol before sex, concurrent sexual partners, men’s circumcision status, whether the participant’s employment type was associated with high risk of acquiring HIV, and whether partner had a high-risk employment type. ART coverage was defined as the proportion of all participants living with HIV who self-reported ART use. Viral suppression was defined as a viral load less than 1000 copies/ml per WHO recommendations [26]. Each individual reported number of sexual partners in the past 12 months and for those who reported more one sexual partner were categorized as having multiple sexual partners. Transactional sex was defined as self-report of having exchanged money, gifts, or favors for sex.

### Statistical analysis

We first conducted descriptive analyses of demographic variables and key outcomes to characterize the sample overall and assess differences between participants who were affected by floods pre- and post-COVID-19 emergence and lake flooding. Direct impacts of household flooding were estimated using difference-in-differences, specifically before-to-after changes in primary outcomes (HIV risk score, ART use, viral load suppression) in flooded and non-flooded households. For each outcome, modified Poisson regression models included indicators for flooding (household flooded; yes or no) and survey period (pre-or post-COVID 19), with difference-in-differences measured through the coefficient of the interaction term between the two variables. Associations were reported were unadjusted and adjusted prevalence ratios (aPR) with 95% confidence intervals (CI). We also conducted a matched-pair-analysis using conditional logistic regression to assess significant changes in primary outcomes over calendar time within individual participants.

### Ethical approval

The RCCS was reviewed and approved by the Research and Ethics Committee at the Uganda Virus Research Institute, and it is registered at the Uganda National Council for Science and Technology. Additional approvals were obtained from the Johns Hopkins Committee on Human Subjects Protections. Community consent was obtained through dialogue meetings with local leaders and the community advisory board prior to the start of each survey visit and sometimes intermediately. Verbal informed consent is provided at household census level by informants or an adult in the household and both verbal and individual written informed consent at the RCCS survey participation level by those eligible and willing. All the datasets were anonymized by the Rakai Health Sciences Program data managers. Permission to use drones in the community was obtained from the local leaders and the Uganda Police.

## RESULTS

In the pre-COVID survey a total of 4,366 were survey eligible, of whom 2,209 participated (50.6%). The primary reason for non-participation was absence for work or school at time of survey, rather than refusal (Supplemental Table 1). Between the pre- and post-COVID-19 surveys, 41.1% (n=855) of pre-COVID-19 participants were lost to follow-up. Direct flooding and COVID-19 impact on primary outcomes were assessed in the remaining 1,226 participants. Out-migration and absence from work were the dominant reasons for lost to follow-up (Supplemental Table 2). Participants in both surveys and included in the final analysis significantly differed from those lost to follow-up on several factors, including sex, age, marital status, occupation, HIV risk score, and purported engagement in transactional sex (Supplemental Table 3).

Of the 1,226 participants in the pre- and post-COVID-19 surveys (Table 1), 506 were in households directly impacted by floods. Of those who were affected by floods, 40.1 % (n=203) were female and 66% (n=334) were married at the post-COVID-19 survey. Demographic characteristics of those whose households were not directly impacted by floods were similar. Compared to the initial survey, participants in post-COVID-19 survey were somewhat older, which was expected because of the closed cohort study design.

**Table 1a:** Sociodemographic characteristics of participants by flooding category in the pre- and post- COVID-19 surveys

|  | **Pre-COVID** |  |  | **Post-COVID** |  |  |
| --- | --- | --- | --- | --- | --- | --- |
| **Demographic characteristics** | **Non-Flooded (N=720)** | **Flooded (N=506)** | **Chi-square p-value** | **Non-Flooded (N=720)** | **Flooded (N=506)** | **Chi-square p-value** |
| **Sex** |  |  |  |  |  |  |
| Female | 310 (43.1) | 203 (40.1) | 0.305 | 309 (42.9) | 203 (40.1) | 0.328 |
| Male | 410 (56.9) | 303 (59.9) |  | 411 (57.1) | 303 (59.9) |  |
| **Age group** |  |  |  |  |  |  |
| 15-24 | 142 (19.7) | 77 (15.2) | 0.154 | 78 (10.8) | 33 (6.5) | 0.036 |
| 25-34 | 310 (43.1) | 216 (42.7) |  | 256 (35.6) | 196 (38.7) |  |
| 35-44 | 237 (32.9) | 190 (37.5) |  | 294 (40.8) | 199 (39.3) |  |
| 45+ | 31 (4.3) | 23 (4.5) |  | 92 (12.8) | 78 (15.4) |  |
| **Marital status** |  |  |  |  |  |  |
| Married | 453 (62.9) | 316 (62.5) |  | 490 (68.1) | 334 (66) |  |
| Separated/Divorced | 182 (25.3) | 141 (27.9) |  | 179 (24.9) | 136 (26.9) |  |
| Never married | 85 (11.8) | 49 (9.7) | 0.370 | 51 (7.1) | 36 (7.1) | 0.720 |
| **Occupation** |  |  |  |  |  |  |
| Agriculture | 49 (6.8) | 41 (8.1) |  | 52 (7.2) | 37 (7.3) |  |
| Administrative | 14 (1.9) | 6 (1.2) |  | 8 (1.1) | 4 (0.8) |  |
| Trading | 175 (24.3) | 100 (19.8) |  | 173 (24) | 112 (22.1) |  |
| Fishing | 226 (31.4) | 195 (38.5) |  | 224 (31.1) | 193 (38.1) |  |
| Student | 16 (2.2) | 6 (1.2) |  | 2 (0.3) | 0 (0.0) |  |
| Unemployed | 64 (8.9) | 37 (7.3) |  | 66 (9.2) | 38 (7.5) |  |
| Others | 176 (24.4) | 121 (23.9) | 0.072 | 195 (27.1) | 122 (24.1) | 0.205 |

Between the pre- and post-COVID-19 surveys, we observed somewhat lower levels of HIV risk behaviors and higher levels of HIV treatment coverage (Supplemental Table 4). For example, we observed significant declines in inconsistent condom use with non-marital partners and self-reported transactional sex, irrespective of household flooding status (Table 1b, Figure 3). Relatedly, we observed a decline in HIV risk score among HIV-seronegative participants, also regardless of flooding status (Figure 4, Supplemental Table 4). There was a statistically significant increase in ART uptake among flooded households (92.0% to 97.9%, p=0.008) and non-flooded households (91.3% to 96.8%, p=0.007). There was a non-statistically significant increase in HIV viral load suppression among flooded households (85.0% to 88%, p=0.108) and non-flooded households (87.9% to 88.8%, p=0.126).

**Table 1b:** Comparison of HIV-associated behavioral characteristics of among 1,266 participants by household flooding status in the pre-COVID-19 and post-COVID-19 surveys.

|  | Flooded (N=506) |  |  | Non-Flooded (N=720) |  |  |
| --- | --- | --- | --- | --- | --- | --- |
| HIV associated behaviors | Pre-COVID | Post-COVID | P value | Pre-COVID | Post-COVID | P value |
| **Number of sexual partners in the past 12 months** |  |  |  |  |  |  |
| 0 to 1 partner | 309 (61.1) | 305 (60.3) |  | 423 (58.8) | 420 (58.3) |  |
| More than 1 partner | 197 (38.9) | 201 (39.7) | 0.797 | 297 (41.3) | 300 (41.7) | 0.873 |
| **Inconsistent condom use with non-marital partners** |  |  |  |  |  |  |
| No | 295 (58.3) | 320 (63.2) |  | 421 (58.5) | 452 (62.8) |  |
| Yes | 211 (41.7) | 186 (36.8) | 0.108 | 299 (41.5) | 268 (37.2) | 0.095 |
| **Transactional sex** |  |  |  |  |  |  |
| No | 365 (72.1) | 388 (76.7) |  | 501 (69.6) | 534 (74.2) |  |
| Yes | 141 (27.9) | 118 (23.3) | 0.098 | 219 (30.4) | 186 (25.8) | 0.053 |
| **Alcohol use in the past 12 months** |  |  |  |  |  |  |
| Yes | 294 (58.1) | 269 (53.2) | 0.114 | 416 (57.8) | 399 (55.4) | 0.366 |
| No | 212 (41.9) | 237 (46.8) |  | 304 (42.2) | 321 (44.6) |  |
| **HIV status** |  |  |  |  |  |  |
| Negative | 319 (63.0) | 314 (62.1) | 0.745 | 455 (63.2) | 443 (61.5) | 0.514 |
| Positive | 187 (37.0) | 192 (37.9) |  | 265 (36.8) | 277 (38.5) |  |
| **ART status** |  |  |  |  |  |  |
| Not on ART | 15 (8.0) | 4 (2.1) | 0.008 | 23 (8.7) | 9 (3.2) | 0.007 |
| Yes | 172 (92.0) | 188 (97.9) |  | 242 (91.3) | 268 (96.8) |  |
| **Viral load suppression** |  |  |  |  |  |  |
| Suppressed | 159 (85.0) | 169 (88.0) | 0.108 | 233 (87.9) | 246 (88.8) | 0.126 |
| Not suppressed | 27 (14.4) | 17 (8.8) |  | 30 (11.3) | 20 (7.3) |  |
| Missing | 1 (0.5) | 6 (3.1) |  | 2 (0.7) | 11 (3.9) |  |

**Figure 3:**
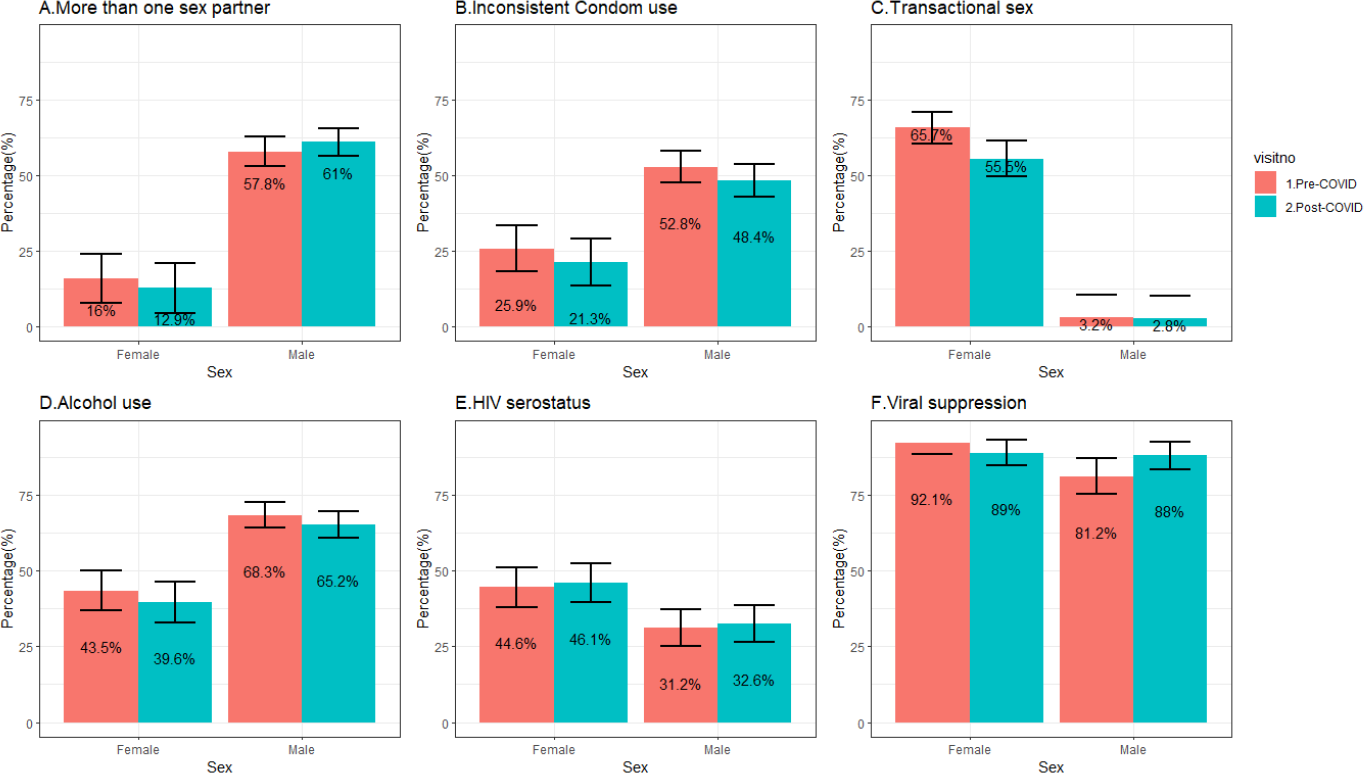
Comparison of HIV risk behaviors and HIV outcomes by sex and COVID exposure.

**Figure 4:**
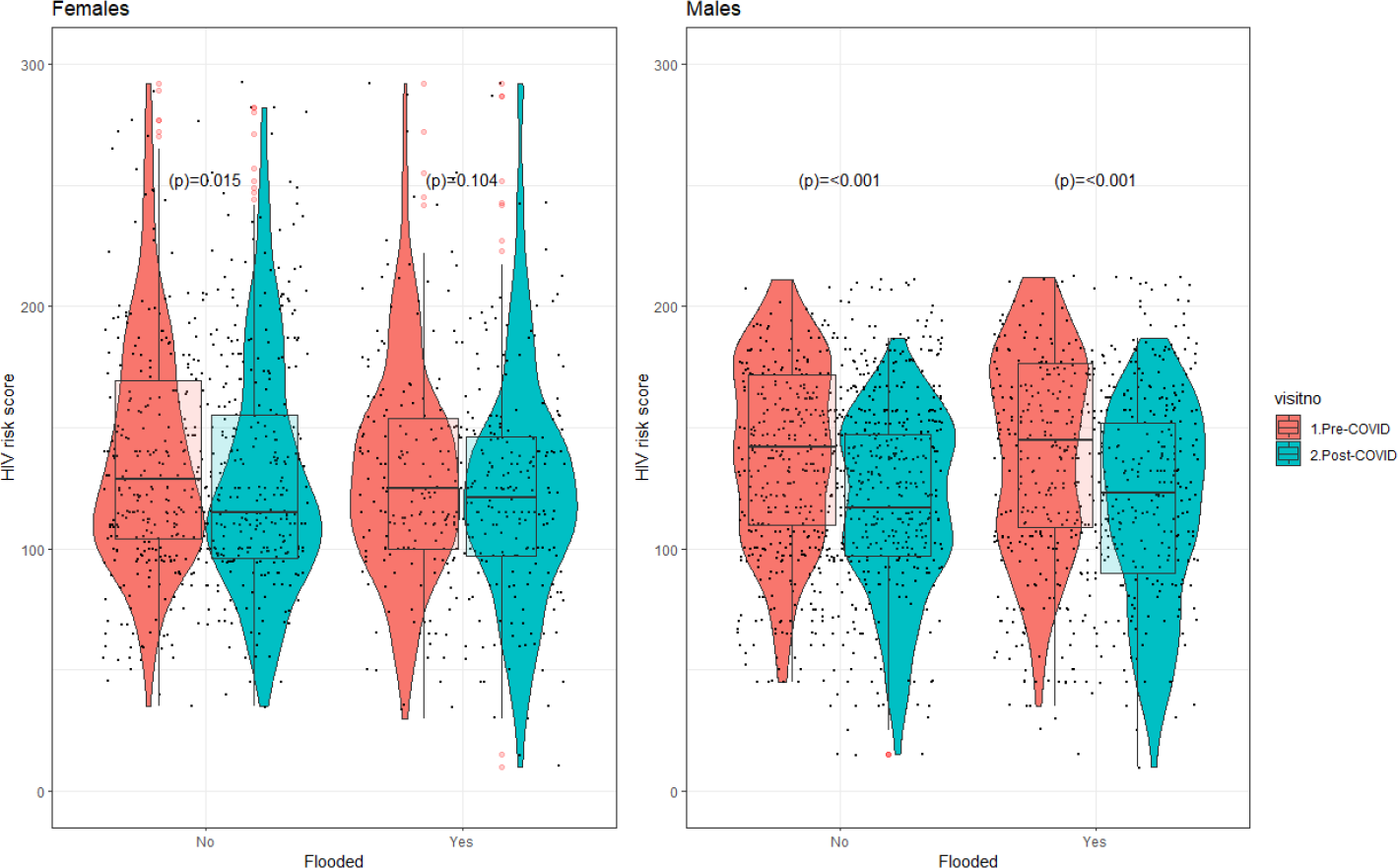
Comparison of overall HIV risk scores among females and males, by exposure to flooding and COVID.

We observed 17% decline in transactional sex (aPR=0.83, 95% CI: 0.75-0.92) and 28% decline in the overall HIV risk score (aPR=0.83, 95% CI: 0.75-0.92) among HIV negatives between the pre- and post-COVID periods. ART coverage increased by 5% (from 91.6% to 97.2%, p<0.001) between the pre- and post-COVID periods among people living with HIV (Table 2).

**Table 2:** Unadjusted and adjusted modified Poisson statistical model for HIV risk behaviors and outcomes in pre- and post-COVID-19 periods.

|  | PR (95% CI) | P value | Adjusted PR (95% CI) | P value |
| --- | --- | --- | --- | --- |
| **Sexual partners in the 12 months** |  |  |  |  |
| Pre-COVID | 1 |  | 1 |  |
| Post-COVID | 1.01 (0.92-1.11) | 0.774 | 1.05 (0.96-1.14) | 0.299 |
| **Inconsistent condom use with non-marital partner** |  |  |  |  |
| Pre-COVID | 1 |  | 1 |  |
| Post-COVID | 0.89 (0.81-0.98) | 0.023 | 0.96 (0.88-1.06) | 0.438 |
| **Transactional sex** |  |  |  |  |
| Pre-COVID | 1 |  | 1 |  |
| Post-COVID | 0.84 (0.74-0.96) | 0.011 | 0.83 (0.75-0.92) | 0.001 |
| **HIV risk score** |  |  |  |  |
| Pre-COVID | 1 |  | 1 |  |
| Post-COVID | 0.60 (0.53-0.68) | <0.001 | 0.72 (0.64-0.80) | <0.001 |
| **HIV prevalence** |  |  |  |  |
| Pre-COVID | 1 |  | 1 |  |
| Post-COVID | 1.04 (0.94-1.15) | 0.479 | 0.93 (0.85-1.03) | 0.177 |
| **ART (HIV positive only)** |  |  |  |  |
| Pre-COVID | 1 |  | 1 |  |
| Post-COVID | 1.06 (1.03-1.10) | <0.001 | 1.05 (1.02-1.08) | 0.002 |
| **Viral load suppression (HIV positive only)** |  |  |  |  |
| Pre-COVID | 1 |  | 1 |  |
| Post-COVID | 1.05 (1.01-1.10) | 0.027 | 1.03 (0.98-1.07) | 0.249 |

In difference-in-difference analyses, we found no significant differences in HIV risk behaviors, disease burden, or viral suppression outcomes, comparing the group affected by floods to those not affected by floods in the pre- and post-COVID time periods (Table 3). Results were similar in matched-pair conditional logistic regression analyses (Supplemental Table 4).

**Table 3:**
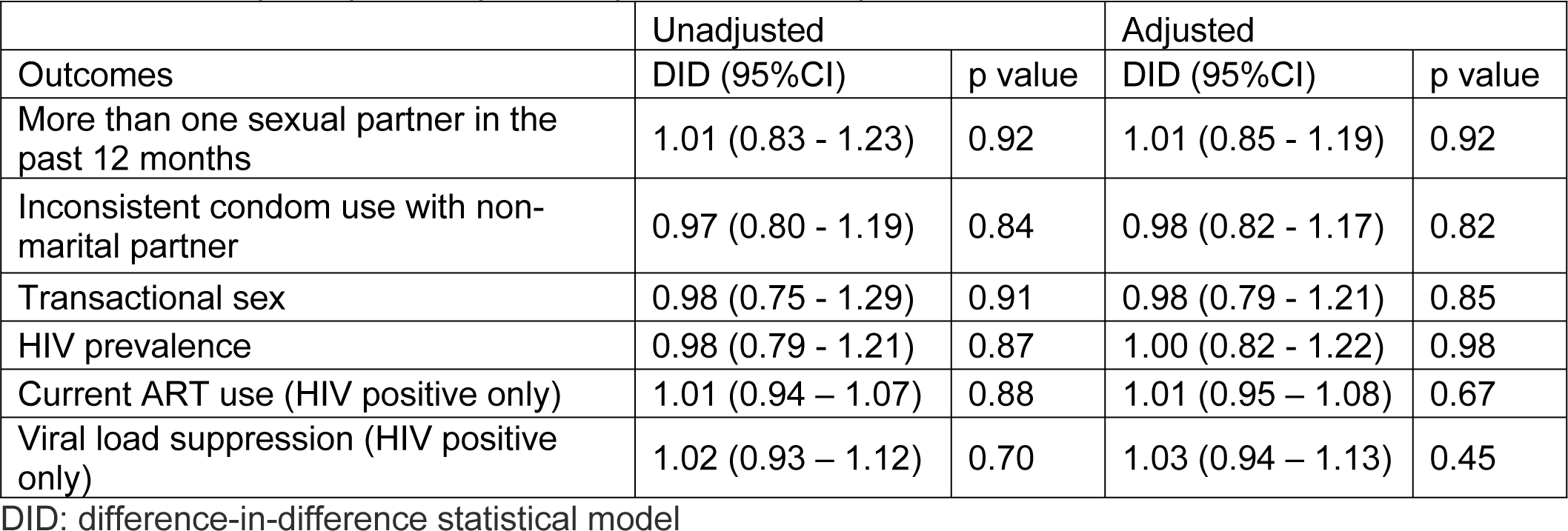
Unadjusted and adjusted difference-in-difference statistical model between flooded and non-flooded participants in pre- and post-COVID-19 period.

| Outcomes | Unadjusted |  | Adjusted |  |
| --- | --- | --- | --- | --- |
|  | DID (95%CI) | p value | DID (95%CI) | p value |
| More than one sexual partner in the past 12 months | 1.01 (0.83 - 1.23) | 0.92 | 1.01 (0.85 - 1.19) | 0.92 |
| Inconsistent condom use with non-marital partner | 0.97 (0.80 - 1.19) | 0.84 | 0.98 (0.82 - 1.17) | 0.82 |
| Transactional sex | 0.98 (0.75 - 1.29) | 0.91 | 0.98 (0.79 - 1.21) | 0.85 |
| HIV prevalence | 0.98 (0.79 - 1.21) | 0.87 | 1.00 (0.82 - 1.22) | 0.98 |
| Current ART use (HIV positive only) | 1.01 (0.94 - 1.07) | 0.88 | 1.01 (0.95 - 1.08) | 0.67 |
| Viral load suppression (HIV positive only) | 1.02 (0.93 - 1.12) | 0.70 | 1.03 (0.94 - 1.13) | 0.45 |

DID: difference-in-difference statistical model

## DISCUSSION

Despite the co-occurrence of flooding and the COVID-19 pandemic in a high HIV risk population, our analysis observed declines in transactional sex and HIV risk score among HIV-negatives as well as an increase in ART coverage. We observed no adverse impact of these extreme environmental conditions on HIV risk behaviors, seroprevalence, or virological outcomes among individuals who remained in the community.

These findings are consistent with findings from a study that assessed effects of COVID on HIV services from eleven sub-Saharan African countries, which noted an increase in the number of people living with HIV who are on ART and virally suppressed during the COVID-19 pandemic [27]. In East Africa, despite significant societal and environment disruptions, relatively few ART adherence challenges were reported [28, 29]. We observed declines in inconsistent condom use with non-marital sexual partners, transactional sex, and overall HIV risk score comparing pre-COVID-19 and post-COVID-19 in both the flooded and non-flooded groups in this hyperendemic fishing community. It is possible that the existence of COVID-19 and related health messages during pandemic response increased overall health consciousness [30, 31]. Notably, a reduction in HIV risk factors was observed prior to the dual crises, consistent with what has been reported in this population between the pre- and post-COVID-19 periods [21]. It is not inconceivable that there was a decline in inconsistent condom use, given that there was a reduction in transactional sex as well. In times of adverse crisis of this nature, it could be that there was little to transact in, therefore transactional sex may not have been an option. Also, disposal income could have been affected so people could hardly engage in luxuries [32]. Although the opposite is possible too (i.e., people are more likely to engage in transactional sex in times of crisis), this is premised on the fact that there is one group that has material items and another that lacks material items. In the case of this community at this time, there is no reason to suspect that some group(s) had preferential access to items that could be used for transactions.

Resilience to flooding and COVID among PLWHIV in this population may have been bolstered by the presence of supportive social networks. People living with HIV who have strong support systems, such as healthcare providers, community peer leaders and friends can rely on these networks for assistance, emotional support, and access to necessary resources during times of emergency [33-35].

In addition, this study focused on individuals who survived life-threatening trauma from flooding and COVID. Experiencing these traumas may lead individuals to have enhanced awareness of their vulnerability and the importance of maintaining good health and prioritizing health care services [36].

The HIV resilience observed in this population may also be attributed to innovative programs like use of motorcycle taxis, multi-month dispensing of ARVs, and ART delivery approaches (e.g. fast-track, home, peer, community pharmacy, and community client-led models) that were implemented during the COVID pandemic period in this region [23]. Addressing HIV program needs could ameliorate the effect of disaster on HIV programming, as has been reported in other similar settings [7].The use of telemedicine, as was the case in this setting, has also been reported to improve health outcomes on HIV risk indicators during COVID-19 [37].

This study had limitations. First, the flooding in Lake Victoria and COVID-19 happened at around the same time, making it difficult to assess the impact of floods or COVID-19 separately. Second, the community experienced other social disruptions: for example, the Ugandan army targeted communities which depended on subsistence fishing on Lake Victoria and other lakes to eliminate what the state considered illegal fishing after a 2020 update in the national fisheries and aquaculture policy [38]. These and other unmeasured activities in fishing communities may make it difficult to control for all confounding factors in assessing the effect of flooding and COVID-19 on HIV risk behaviors.

In conclusion, despite a high background burden of HIV, the COVID-19 pandemic, and serious flooding, we observed no adverse impact on HIV risk behaviors, burden, or virological outcomes in this HIV hyperendemic fishing community. These findings suggest that innovative programs and population resilience may have sustained good HIV outcomes. Understanding exactly what HIV programs and personal/community-level strategies worked to maintain good public health outcomes despite extreme environmental and pandemic conditions may help improve population health.

## Data Availability

All relevant data are within the paper and its supporting information files. Additional information may be requested from the corresponding author.

## Author contributions

Hadijja Nakawooya – Conceptualization, Data curation, Formal analysis, Methodology, Writing – Original Draft Preparation, Writing – Review & Editing

Victor Ssempijja – Conceptualization, Formal analysis, Writing – Review & Editing Anthony Ndyanabo - Data curation, Formal analysis, Writing – Review & Editing

Ping Teresa Yeh - Investigation, Visualization, Writing – Original Draft Preparation, Writing – Review & Editing

Larry W. Chang - Conceptualization, Writing – Original Draft Preparation, Writing – Review & Editing

Maria J. Wawer - Writing – Review & Editing

Fred Nalugoda -

David Serwadda -

Ronald H. Gray - Writing – Review & Editing

Joseph Kagaayi -

Steven J. Reynolds –

Tom Lutalo –

Godfrey Kigozi -

M. Kate Grabowski – Conceptualization, Methodology, Writing – Original Draft Preparation, Writing – Review & Editing

Robert Ssekubugu – Conceptualization, Investigation, Writing – Review & Editing

## Acknowledgements

We thank the Rakai Community Cohort Study participants and the Rakai Health Sciences Program team for making this study possible.

## Funding

The Rakai Community Cohort Study rounds 19 and 20 were supported by the National Institute of Mental Health (R01MH099733, R01MH105313, R01MH107275, R01MH115799), National Institute of Allergy and Infectious Diseases (R01AI114438, K25AI114461, R01AI123002, K01AI125086, R01AI128779, R01AI143333, R21AI145682, R01AI155080), National Institute on Alcohol Abuse and Alcoholism (K01AA024068), Eunice Kennedy Shriver National Institute of Child Health and Human Development (R01HD091003), National Heart, Lung, and Blood Institute (R01HL152813), and the Bill and Melinda Gates Foundation (OPP1175094). The study was also supported in part by the Division of Intramural Research, National Institute of Allergy and Infectious Diseases. HN received training and support from National Institutes of Health Fogarty International Center (D43TW010557). The funders were not involved in the design of the study and collection, analysis, and interpretation of data or in writing the manuscript.

## Competing interests

The authors declare no competing interests.

## Notes

### Competing Interest Statement

The authors have declared no competing interest.

